# Pooling for SARS-CoV2 Surveillance: Validation and Strategy for Implementation in K-12 Schools

**DOI:** 10.1101/2020.12.16.20248353

**Authors:** Alexandra M. Simas, Jimmy W. Crott, Chris Sedore, Augusta Rohrbach, Anthony P. Monaco, Stacey B. Gabriel, Niall Lennon, Brendan Blumenstiel, Caroline A. Genco

## Abstract

Repeated testing of a population is critical for limiting the spread of the SARS-CoV-2 virus and for the safe reopening of educational institutions such as K-12 schools and colleges. Many screening efforts utilize the CDC RT-PCR based assay which targets two regions of the novel Coronavirus nucleocapsid gene. The standard approach of testing each person individually, however, poses a financial burden to these institutions and is therefore a barrier to using testing for re-opening. Pooling samples from multiple individuals into a single test is an attractive alternate approach that promises significant cost savings - however the of specificity and sensitivity of such approaches needs to be assessed prior to deployment. To this end, we conducted a pilot study to evaluate the feasibility of analyzing samples in pools of eight by the established RT-PCR assay. Participants (1,576) were recruited from amongst the Tufts University community undergoing regular screening. Each volunteer provided two swabs, one analyzed separately and the other in a pool of eight. Because the positivity rate was very low, we spiked approximately half of the pools with laboratory-generated swabs produced from known positive cases outside the Tufts testing program. The results of pooled tests had 100% correspondence with those of their respective individual tests. We conclude that pooling eight samples does not negatively impact the specificity or sensitivity of the RT-PCR assay and suggest that his approach can be utilized by institutions seeking to reduce surveillance costs.

## Introduction

Shutdowns resulting from an effort to control the ongoing COVID-19 pandemic have had a detrimental effect on the education of K-12 school children, especially those in at-risk socioeconomic groups and those with special needs (Loades *et al*. 2020; Masonbrink & Hurley 2020). For example, clinicians note significant weight gains and precipitation of anxiety disorders during COVID-19 (Rundle *et al*. 2020). Educational attainment is an important predictor of future health, mental state, and socio-economic outcomes (Dewalt *et al*. 2004). Returning these children to school in a safe and affordable way is a high priority. In addition to appropriate safety precautions like social distancing and mask wearing, testing of all individuals in a population at regular intervals can reduce transmission via early detection of asymptomatic or pre-symptomatic infectious viral carriers who can be a source of transmission.

Along with limited laboratory equipment, reagents, and resources, the cost of repeated individual testing presents a substantial challenge. One potentially more efficient testing strategy is sample pooling, a broadly developed mathematics subfield wherein samples from individuals are grouped together and tested as a single unit with a single output. Pooled testing can reduce the number of tests performed by a factor of the pool size, reducing cost as well as laboratory throughput demands. The simplest form of pooling is known as Dorfman or two-stage hierarchical pooling (Dorfman 1943). By this method, each pool contains a set number of samples. Each sample is tested once as part of a pool, and again as an individual test only if the pool tests positive. Other, more complex methods assign a single sample to more than one pool to better predict positive samples, thus reducing the number of individual retests necessary (McDermott *et al*. 2020; Mutesa *et al*. 2020; Shental *et al*. 2020). Dorfman pooling for SARS-CoV-2 RT-PCR testing has been shown to be effective at the level of eight samples per group (Barak *et al*. 2020).

The CDC-approved, commonly used diagnostic test for SARS-CoV-2 infection employs reverse transcription-polymerase chain reaction (RT-PCR) to detect viral RNA present in mucosal samples immersed in a liquid buffer (CDC 2020). Sample pooling has been proposed and validated by a number of groups, with the pooling step performed at various points in the diagnostic workflow e.g. 1) extracting RNA from multiple swabs in a single container, 2) combining extracted RNA into a single container for cDNA synthesis, or 3) pooling the cDNA after production on an individual basis. Benefits of pooling later in the process include equal amounts of input per subject and rapid retesting of positive pools if individual samples are stored. However, the earlier pooling occurs in the process the greater cost-savings in reagents. Additionally, direct pooling of individual swabs into a single tube can be done at the site of testing, effectively saving time, labor, and materials. Because of the financial benefits, the Food and Drug Administration (FDA) has already issued the first Emergency Use Authorization (EUA) for pooled testing of SARS-CoV-2 for pools of up to 4 samples (US Food and Drug Administration 2020b).

Recently, the Broad Institute has established a CLIA-approved laboratory with a protocol for performing RT-PCR from RNA extracted from anterior nares dry swabs. Here, we present a proof-of-concept pilot study intended to validate the efficacy of pooled SARS-CoV-2 testing using dry swabs when compared with individual testing.

## Methods

This study was reviewed and approved by the Institutional Review Board of Tufts University (STUDY00000979: Pooled Testing). Eligibility criteria included being a member of the Tufts University faculty, staff, or student populations as outlined by risk of exposure to COVID-19 (https://coronavirus.tufts.edu/testing-at-tufts), ages 18-100.

Tufts University has implemented a comprehensive COVID-19 testing program to enable the safe return of students to campus. Students, faculty, staff and researchers are required to test 1-2 times per week depending on their residential arrangement, frequency and nature of campus use, interactions with each other, and exposure to patients or the general public.

For this program, individuals report to various testing locations and self-swab their anterior nares with a single sterile soft-tip swab (Puritan Medical Products LLC) which is then deposited into a prelabelled plain vacutainer (Beckton Dickinson). Samples are couriered to the Broad Institute 4-6 times per day and analyzed by the established CDC RT-PCR assay with fluorescent detection (FAM-labelled probes) (US Food and Drug Administration 2020a).

In September and October of 2020 we obtained samples from 2,032 members of the Tufts University community undergoing routine screening. Individuals were not restricted from participating in the study multiple times. Upon arrival at the testing site, participants were provided with two swabs and instructed to swab one per nostril and deposit each in a separate vacutainer. The method of collection was the dry swab method described here (US Food and Drug Administration 2020a). At the end of each study day, one sample per individual was sent for regular individual testing while the other was processed for pooled testing. For pooling, individual samples were removed from their vacutainer and placed in a 50 mL Falcon tube (Thermo Fischer Scientific). One hundred and fifty one pools were processed as groups of eight samples. Eleven pools were made up of seven community samples plus an additional sample obtained from an individual under investigation based on a previous positive SARS-CoV-2 RT-PCR test. Of these eleven samples, only two were positive in this study. The remaining nine were likely cases of prior infection. Another one hundred and twelve pools were made up of seven community samples plus an eighth sample prepared by the Broad Institute as described in their EUA (US Food and Drug Administration 2020a). Briefly, these samples were generated by resuspending nasal swab material from known positive cases and spraying it on to new swabs as previously described (Cleary *et al*. 2020). For RNA extraction and RT-PCR, pooled samples were subjected to the same conditions as individual samples, aside from an increased resuspension volume at the start. All samples were analyzed within 24 hours of collection.

## Results

For this study, we selected eight samples per pool for financial and logistical reasons. Financially, the cost and efficiency of various pooling ratios changes as a function of the percent positivity in the population (**Figure 1**). A pool size of eight was the most cost effective per person for positivity rates ranging from 2.22% to 2.86%. At positivity rates lower than 2.22% it is still cost-effective when compared with pools of 9-10 (at a positivity rate of 0.00%, a pool of 8 costs $5.00/person while a pool of 10 costs $4.00/person). Large pool sizes are more efficient and cost effective at lower community positivity rates, but the larger the pool, the more rapidly it increased the cost as the percent positivity increased (**Figure 1**). It is critical that the pool size is flexible to accommodate varying positivity rates in the population. To ease scalability and adoption, it was also important in this study to 1) minimize the operational transition from individual to pooled testing, and 2) pool samples at the site of collection using standardly available materials. Because of these factors, including limitations of rehydration volumes and tube sizes, we did not consider pools larger than 10.

**Figure 1.**
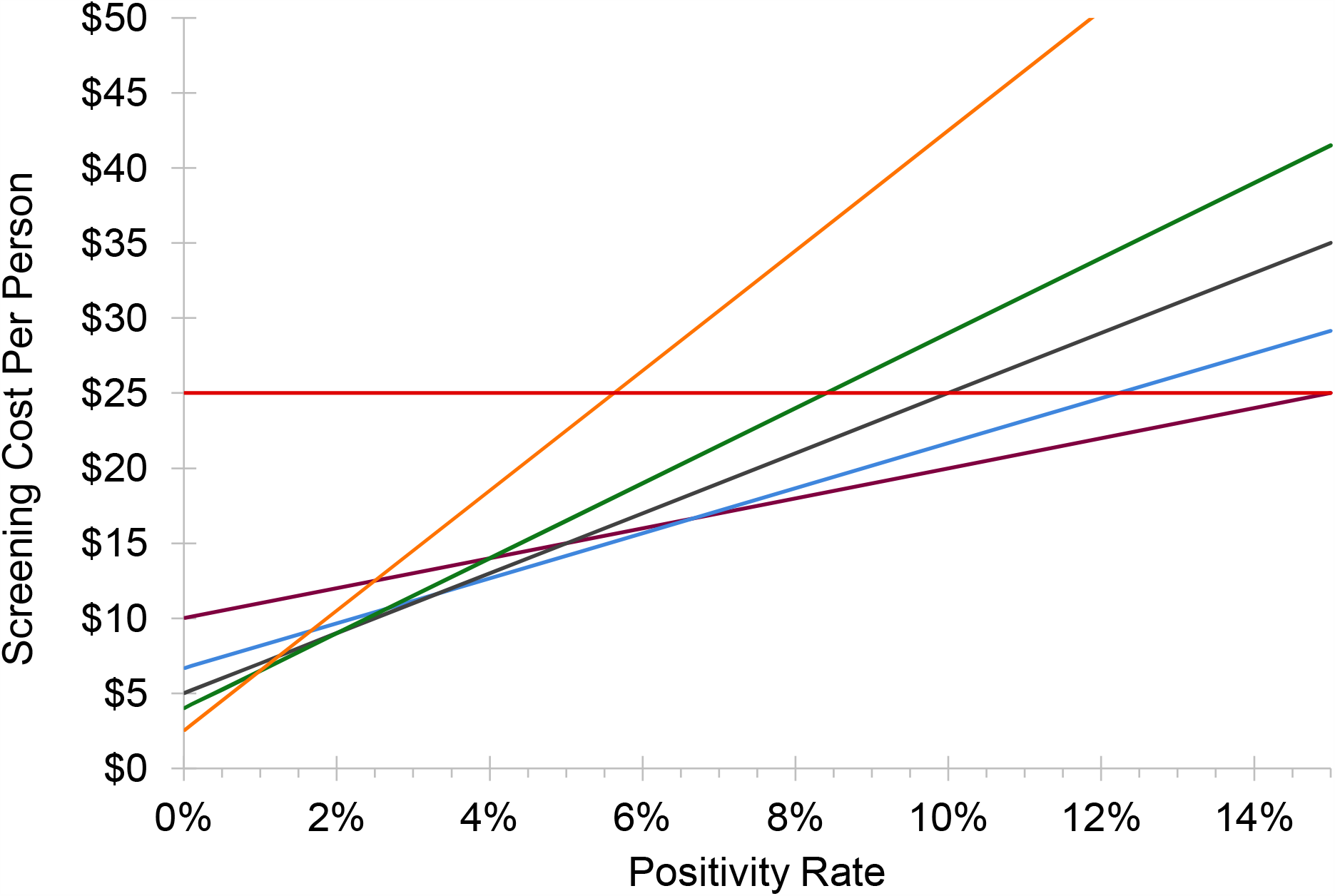
Cost of individual versus pooled testing. Cost per person of testing as a function of the percent positivity of the population tested. This model assumes an individual test cost of $25 and a pooled test cost of $40 due to additional processing steps required. Red line: individual testing, maroon line: pool size of 4, blue line: pool size of 6, grey line: pool size of 8, green line: pool size of 10, orange line: pool size of 16. Equation for pooled testing: ($40/pool size) + (% positivity x pool size x $25).

Over three weeks we collected 2,032 pairs of samples from 1,576 students, faculty, and staff already subject to regular COVID-19 surveillance testing at Tufts University (**Figure 2**). Of these, 1,973 samples came back negative (**Figure 3**). Of the eleven samples taken from individuals under investigation due to possible SARS-CoV-2 exposure, only a single individual was responsible for the two positive samples from our community-based sampling. The study was originally designed to assess sensitivity by collecting only natural positive cases. However, the very low positivity rate within the Tufts community made it impossible to obtain our target number of pools with at least one positive within a reasonable period of time. Thus, to obtain sufficient positive pools we implemented the artificial spiking protocol described in the methods, wherein pools of seven swabs obtained from community members were supplemented with an additional swab generated from known positive samples prior to RNA extraction. In testing individual samples, we observed a collection failure rate of 2.36% (48 samples) (**Figure 3**). Of these, 24 samples were discarded due to a single laboratory cataloging error, four were lost on site, eleven were discarded due to improper collection methods (excessive material on the swab, swab inverted, or contamination with blood), and nine produced an invalid RT-PCR result, likely due to a lack of RNA present. For unknown reasons, one of the laboratory-generated spiked samples also produced an invalid RT-PCR result.

**Figure 2.**
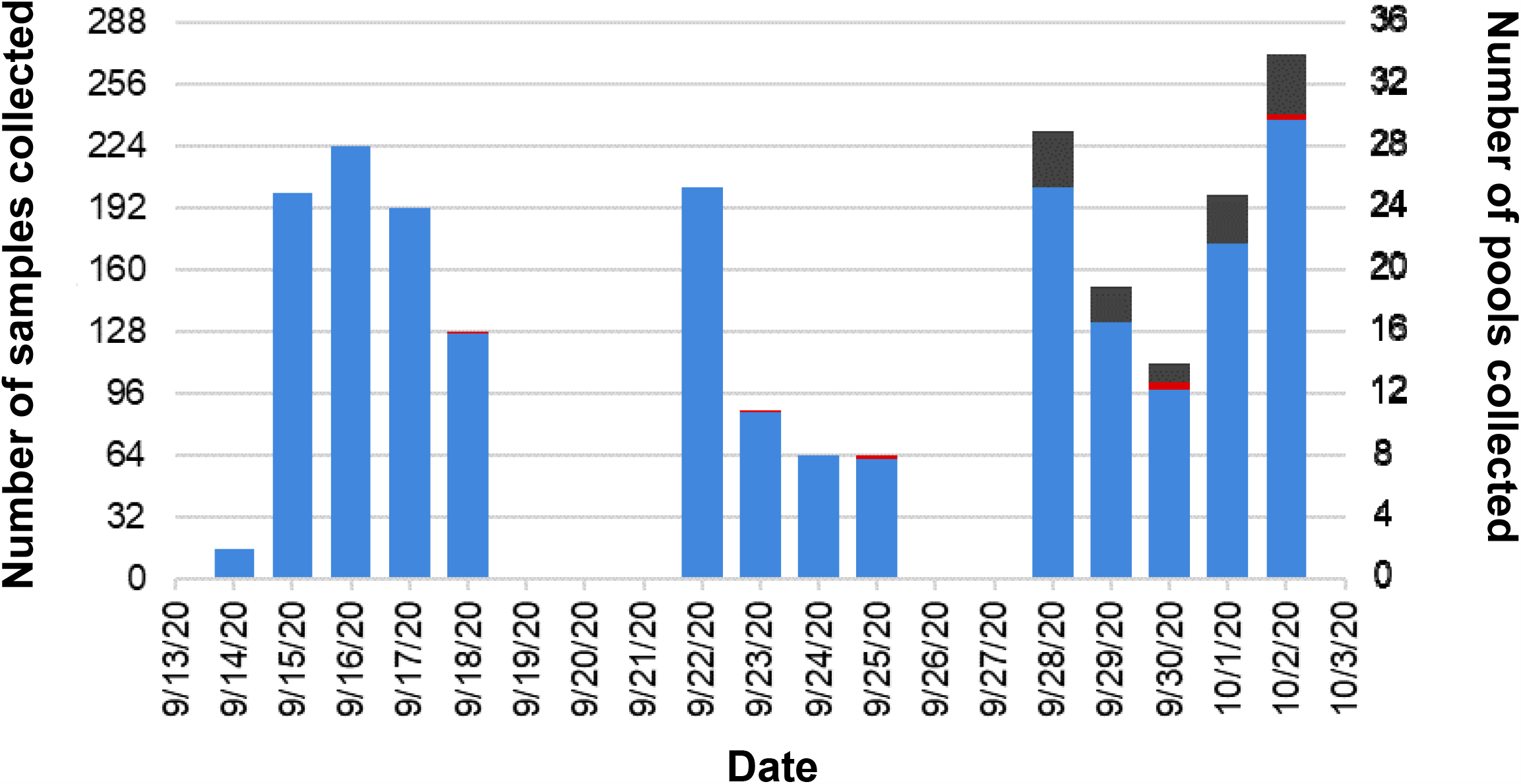
Pooled Pilot Study Timeline. Two thousand and thirty-two pairs of samples were collected over a three-week period. Two thousand and twenty-one of those pairs were collected from the Tufts University community (blue). The other 11 pairs were collected from individuals under investigation and in quarantine due to possible exposure to SARS-CoV-2 (red). During the third week of the study, the low positivity rate in the Tufts community necessitated the inclusion of laboratory-generated positive samples (grey).

**Figure 3.**
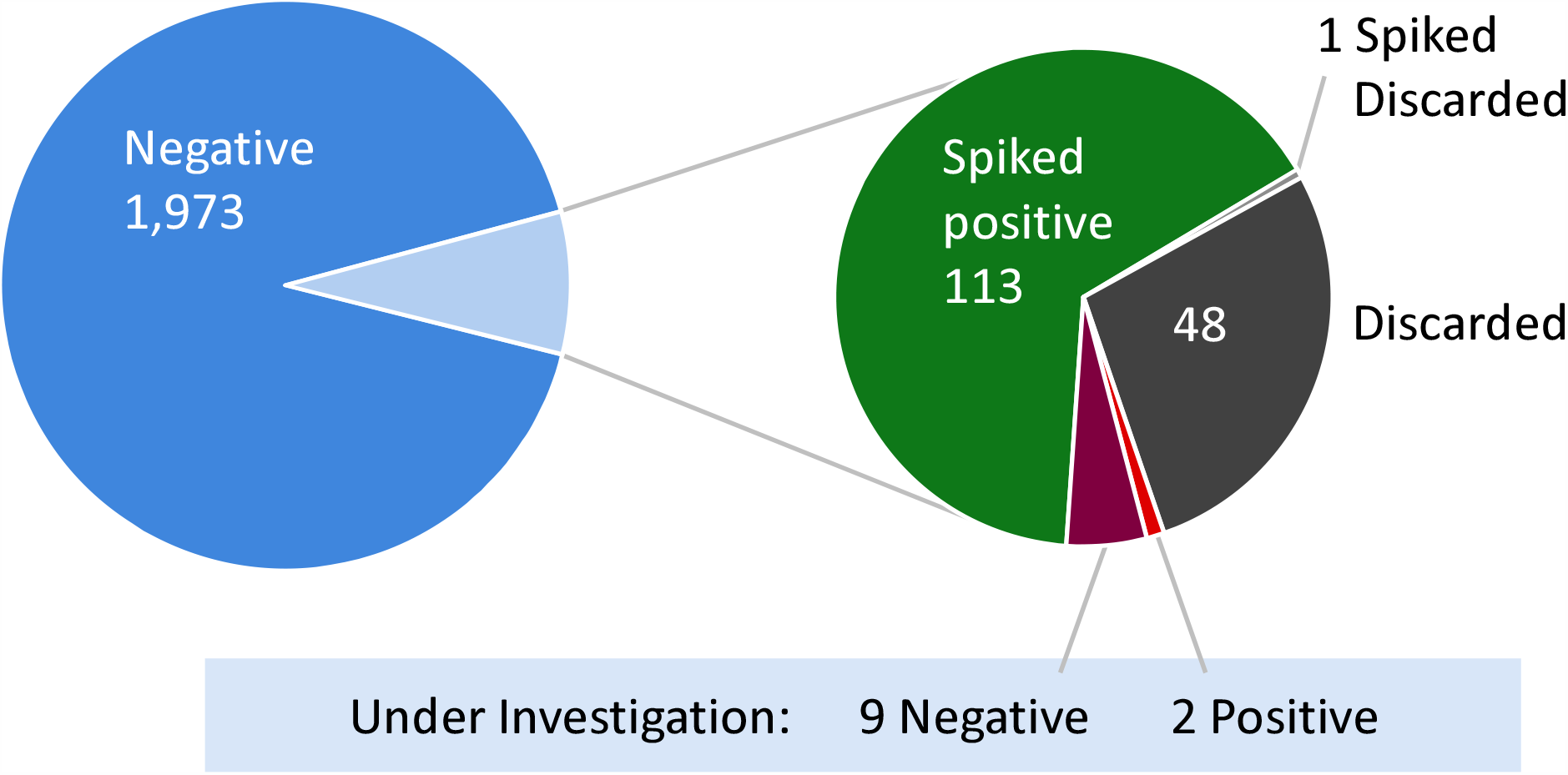
Individual sample results of SARS-CoV-2 RT-PCR testing. Of the 2,032 community-sourced and 114 laboratory-generated pairs of samples, one sample from each pair was tested as an individual using the established SARS-CoV-2 RT-PCR testing protocol. Eleven of the sample pairs were collected from individuals under investigation and in quarantine due to possible exposure to SARS-CoV-2 (red and maroon).

To evaluate the efficacy of pooling samples, results of the individual samples were compared with results of their respective pools (**Table 1**). While 272 sample pools were prepared and successfully analyzed, data from 32 pools was discarded due to a failed individual assay for one or more of the paired samples in the pool as described above. Thus, data for a total of 240 pools were included - 133 with samples of unknown SARS-CoV-2 status and 107 spiked with a known positive. We observed 100% congruency between the approaches; all pools containing a swab whose individual counterpart had tested positive also tested positive and all pools for which individual samples were all negative also tested negative.

**Table 1.**
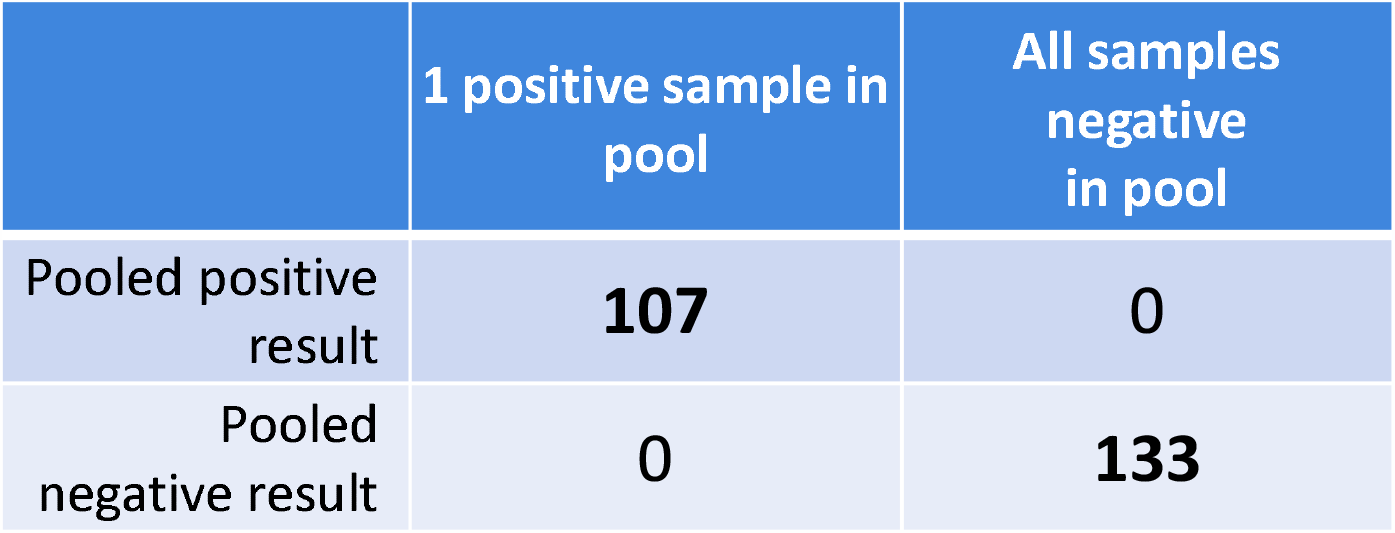
Comparison of pooled with individual sample results. 272 pools were tested. 32 pools were discarded because one or more of the corresponding individual sample tests was invalid, leaving 240 pools which corresponding exactly with the individual results.

It has been suggested that pooling samples may dilute viral material, since every negative sample added will dilute the pool when compared with a single positive sample e.g. a pool containing one positive sample and seven negative samples will be diluted by a factor of eight. Since reaction products of RT-PCR double every cycle, we would expect an eight-fold dilution to result in an addition of three (?cycles) to the Ct when compared with the original positive sample. To evaluate the effect of pooling on PCR amplification cycles, we compared the Ct value of positive pools to those of the corresponding individual sample PCR result (**Figure 4**). We observed little correlation between the N1 or N2 Ct values for individual samples and the comparable Ct for the corresponding pool. We did observe that the pool Ct values were slightly higher than the individual Ct values, as was expected when measuring a single positive sample versus a pool consisting of a single positive sample and 7 negative samples. To demonstrate that the differences in Ct were not associated with the total amount of genetic material in the pool, we also compared the positive individual and its positive pool Ct values for human RNase P gene (RP), an indicator of human genetic content.

## Discussion

Pooled sample testing is an effective strategy to reduce the cost of regular screening testing during pandemics. We conducted a pilot study to evaluate the validity of pooling anterior nasal swabs for the detection of SARS-CoV-2 by RT-PCR. While others have validated pooling of “wet swabs” (de Salazar *et al*. 2020; Gupta *et al*. 2020; Kim *et al*. 2020b), this is the first evaluation of pooled testing using dry swabs. Dry swabbing presents a substantial benefit over wet by saving reagents, thus saving money as well as reducing vulnerability to the supply-chain issues that have interfered with testing capacity during the COVID-19 pandemic.

Potential concerns with pooled testing include the dilution of sample resulting in false negatives and the accumulation of contaminants resulting in false positives. We observed a 100% congruence between the results of the pooled and individual analyses – indicating that there is no loss of specificity or sensitivity when performing SARS-CoV-2 screening from pools of eight dry swabs compared to individual analyses. Given that the clinical specificity and sensitivity of the CDC’s 2019-nCoV Real-Time RT-PCR Diagnostic Panel are 100% (13/13; 95% CI: 77.2%– 100%) and 100% (104/104; 95% CI: 96.4%–100%) (CDC 2020), respectively, this provides great confidence in identifying all positive individuals using pooling. A recent publication by members of the Broad Institute established a reduction in sensitivity that is roughly linear with the log of the dilution factor employed by simulating pooling under varying population prevalence, pool size, and population size (Cleary *et al*. 2020). Specific recommendations for pooled testing from the FDA have only recently become available (US Food and Drug Administration 2020b), and further investigation is necessary to evaluate its efficacy for specific circumstances. For this study, we selected eight samples per pool for logistical and financial reasons described above. Our protocol has a ceiling of ten samples because it calls for rehydrating swabs as a pool. This minimizes labor costs since all subsequent steps can follow the established semi-automatic individual testing workflow. Other groups have successfully performed pooling with different pool numbers (de Salazar *et al*. 2020; Gupta *et al*. 2020; Kim *et al*. 2020b; McDermott *et al*. 2020; Mutesa *et al*. 2020; Shental *et al*. 2020), and conceivably other protocols where this is not a barrier could conceivably take advantage of larger pools. However, as we have shown the cost of large pool sizes increases rapidly with percent positivity. A far more cost-effective approach would be to have a three-tiered system (Black *et al*. 2015), but that would require more labor and/or a customized automated workflow.

One clear drawback of pooled testing is that all individuals in positive pools need to be retested to identify the positive individual. A few approaches can be used to facilitate this: individuals from the positive pools can be asked to return for retesting or every individual provides two samples with the second stored for retesting. The former approach places the burden on individuals to return for retesting and quarantine before their second individual result is obtained. Moreover, it may pose additional risk from spreading as infectious individuals return to testing sites. The latter approach places the burden on the screening organization as they would need to store and retrieve samples as well as use additional supplies to collect those samples. Because of the logistical burden and wastage of supplies we prefer the retesting approach. While the CDC RT-PCR test is the current gold standard, some antigen tests currently under development have promise as an alternative. If they can achieve the same sensitivity and specificity as RT-PCR, the exact same pooled testing model could be used with antigen tests instead, if they are cheaper or more rapid.

We performed a cost-benefit analysis to better understand the potential savings that could be realized through pooled testing. Although fewer samples are processed, additional handling is required to prepare and process the pooled samples. Based on a sample pool size of eight and costs of $25 and $40 for individual and pooled tests, respectively, we estimate that savings from using a pooled method of testing versus individual testing would exist for positivity rates under 10% ($19.88 in savings at 0.01% positivity).

It is worth noting that our observation of a low rate of improper test collection (absence of RNA on the swab, excessive material on the swab, swab inverted) among volunteers who collected their own samples (self-swab). In settings where a trained professional is collecting samples, we expect the rate to decrease dramatically. However, when individuals self-swab it will be impossible to detect individuals with repeat errors in collection methods e.g. those who are not swabbing appropriately because the other samples in the pool will mask it. Therefore, education of self-swabbing subjects will be critical.

A lack of in-person education deprives children of educational and well as social development (Dove *et al*. 2020; Masonbrink & Hurley 2020). Recent meta-analyses suggest that schools, especially PK-8, are not major sources of outbreaks (Dove *et al*. 2020; Kim *et al*. 2020a; National Collaborating Centre for Methods and Tools 2020; Posfay-Barbe *et al*. 2020). Together, these factors provide a compelling argument for establishing in-person learning, even when local population positivity rates are high enough to warrant closing of other social spaces such as restaurants. Maintaining in-person community activities such as K-12 school attendance during the COVID-19 pandemic will rely on early identification and containment of infectious individuals. The pooling method we present here is a simple, scalable way to reduce the cost of regular surveillance screening for such groups.

## Grants

This project was funded by internal funds from the Office of the Vice Provost of Research.

## Data Availability

All data is available upon request.

## Acknowledgements

We would like to thank all the Tufts University students, faculty, and staff who participated in this study; Marie Ellen Caggiano, M.D. for acting as the provider for positive cases; the Armstrong Ambulance Service and their EMTs; Adam Cotton and the Tufts Conference Services; Melanie Marketon and Sarah Augood; Stacey Gabriel, Michelle Cipicchio, Nicholas Fitzgerald, and Doreen Leupold-Mitchell from the Broad Institute.

---

**Figure 5.**
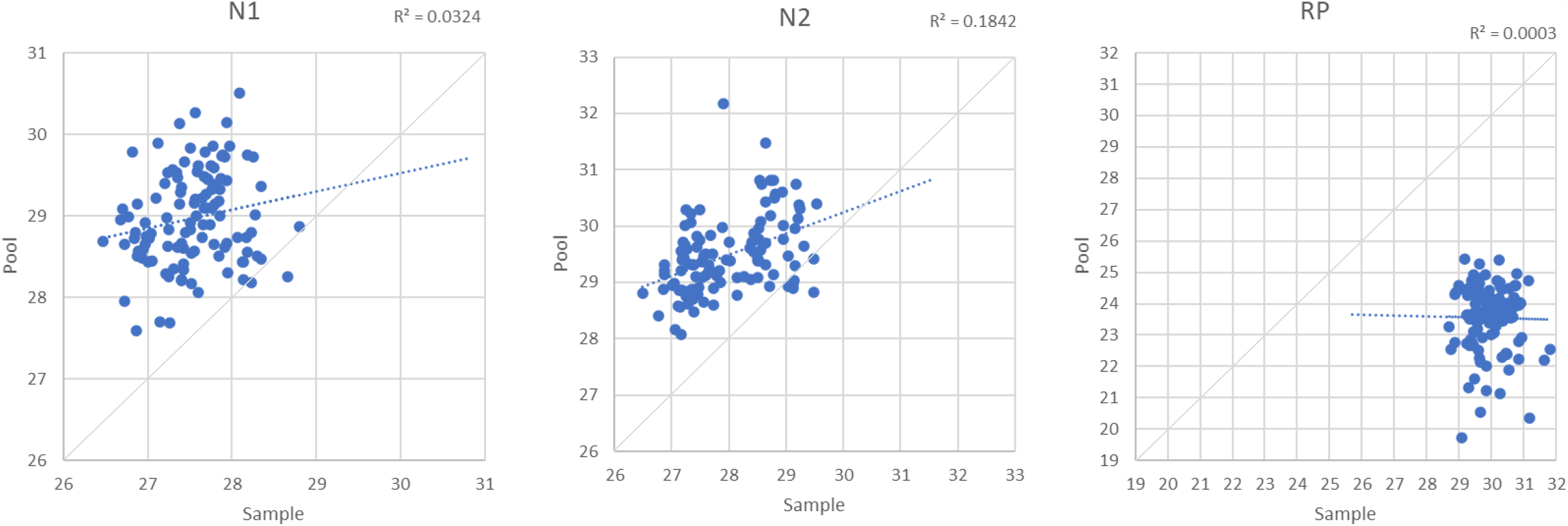
Ct results comparing pool Cts (y-axis) with the Ct value of the corresponding single positive sample (x-axis). Each graph represents a different probe; N1 and N2 detect regions of SARS-CoV-2 virus RNA while RP represents host genetic material abundance. Blue line: trendline of data; grey line: y=x bisector.

## Notes

### Competing Interest Statement

The authors have declared no competing interest.

### Author Declarations

This study was reviewed and approved by the Institutional Review Board of Tufts University (STUDY00000979: Pooled Testing).

